# Inter-relationship of Retinal, Choroidal, and Scleral Thickness in High Myopia

**DOI:** 10.64898/2026.05.13.26353083

**Authors:** Swati Panigrahi, Rohit Dhakal, Kiran K. Vupparaboina, Pavan K. Verkicharla

## Abstract

**Purpose:** Considering that myopia is associated with thinning of the ocular coats, this study investigated the inter-relationship of retinal, choroidal and scleral thickness in foveal regions in Indian high myopes.

**Methods:** A total of 23 high myopes (spherical equivalent refraction ≤-6.00D) aged 16 to 35 years underwent posterior segment imaging with swept-source optical coherence tomography. The retinal, choroidal and scleral thickness was determined using semi-automated custom-designed software at sub-foveal regions. Axial length was determined using Lenstar LS 900 non-contact biometer.

**Results:** The mean ± SD axial length was 30.17 ± 2.23 mm, sub-foveal retinal thickness was 245 ± 28 µm, sub-foveal choroidal thickness was 82 ± 46 µm, and sub-foveal scleral thickness was 254 ± 68 µm. The choroid was significantly thinner compared to the retina and sclera (p<0.001). With a 1 mm increase in axial length, there was no significant variation in sub-foveal retinal (increased by 0.86 µm) and scleral thickness (decreased by 4.31 µm, p≥0.05), but sub-foveal choroidal thickness decreased by 10.35 µm (p=0.02). For a 1D decrease in spherical equivalent refraction, the choroidal thickness reduced significantly (decreased by 5.88 µm, p<0.001), while there was no significant variation in retinal (decreased by 0.68 µm, p=0.55) and scleral thickness (increased by 0.13 µm, p=0.98). The association of the sub-foveal retinal, choroidal, and scleral thickness was weak and was not significant in high myopes (p≥0.10).

**Conclusions:** With increasing axial length and severity of myopia in high myopes, compared to scleral and retinal thickness, the choroidal thickness alone decreased significantly. Our findings indicate that the changes in the choroid do not necessarily reflect the changes in retinal and scleral thickness and highlight the importance of the choroid as a marker for axial elongation even in high myopes.

## Introduction

High myopia is known to be associated with thinning of the outer coats of the eyeball.^1^ Various hypotheses have been proposed to describe cross-talk among the retina, choroid, and sclera during ocular expansion.^2-4^ While there is conflicting evidence on the relation of foveal thickness with refractive error or axial length (both thinning and thickening),^5-12^ a consistent choroidal and scleral thinning has been reported with an increasing degree of myopia.^10, 11, 13, 14^ Further, in an animal experiment, the posterior scleral thickness was found to be significantly correlated with choroidal thickness and choroidal vascularity index under lens-induced myopia.^15^ However, such evidence in humans remains mixed and inconclusive. While Park et al.^16^ reported a positive correlation between choroidal and scleral thickness (sclera thinning with choroidal thinning), other studies reported no or weak association in both pathologic and non-pathologic myopes.^11, 17-19^ It is important to note that previous studies (except Deng et al.^11^) on humans have typically examined the relationship between myopia and thickness of individual layers of the eye (for example, retinal and/or choroidal or scleral thickness), but not the thickness profile of all the three layers simultaneously within the same cohort.^5-14, 20, 21^ Deng et al. investigated retinal, choroidal and scleral thickness in Chinese children that included high myopic eyes up to -11.38 D with a mean refractive error of −6.54 ± 1.35 D.^11^ The findings revealed that the sub-foveal choroid and sclera may get thinner with increasing magnitude of myopia, with no changes in sub-foveal retinal thickness.^11^ That said, it is not clear if this relationship between the three layers continues even in higher forms of myopia. Given the possibility of attaining ceiling effect in biology, we attempted to explore the inter-relationship between sub-foveal retinal, choroidal, and scleral thickness in high myopic eyes with refractive error beyond -6.00 D (up to -26.25 D). Additionally, we investigated the associations of retinal, choroidal, and scleral thicknesses with axial length to better understand the influence of axial elongation on the posterior layers in non-pathologic high myopes. To conduct this observational study, we utilized the data from our previously conducted research work,^21^ in which posterior scleral thickness in myopes was analyzed, and extended it by performing additional analyses to determine corresponding sub-foveal retinal and choroidal thickness.

## Methods

For this study, data were analysed from total of 23 participants aged 16 to 35 years (15 male and 8 female) who were recruited from the comprehensive clinic of L V Prasad Eye Institute, India. The study was approved by the Institutional Review Board of L V Prasad Eye Institute (LEC 09-17-084) and conducted in accordance with the tenets of the Declaration of Helsinki (2013). Written and verbal consent were obtained from all the participants before commencing the measurements. For two participants aged less than 18 years, parents/guardians have provided the written consent, while the children gave the verbal assent. To avoid the involvement of pathologic changes in thickness analysis, all individuals who had high myopia without any abnormalities in the OCT B-scan were included. Any other ocular or systemic condition that may influence refractive error was excluded. Before measurements, a comprehensive eye examination was performed for all the participants, along with dilated fundus evaluation. Pupillary dilation was achieved by instilling two drops of Itrop plus (phenylephrine 5% wt/vol and tropicamide 0.8% wt/vol; Cipla Ltd., Mumbai, India) with each drop instilled in 5-minute interval to achieve maximum pupillary dilation. An average of three consecutive measurements of axial length with a Lenstar LS 900 optical noncontact biometer (Haag Streit, Köniz, Switzerland) and average of five central refraction values determined using a Shin-Nippon open field autorefractor (Nvision-K 5001, Tokyo, Japan) with the target placed at 3 meters from the eye was considered for analysis. For posterior segment imaging of retina, choroid and sclera, images were obtained utilising Topcon-3 DRI Triton SS-OCT (Tokyo, Japan). The SS-OCT with enhanced depth imaging mode (EDI) provides higher penetration up to the choroid and sclera.^22^ All measurements were conducted by a single experienced examiner under normal room illumination (<250 lux), and only right eye OCT images were considered for analysis. Raw B-scan OCT images along the naso-temporal meridian were acquired using a 12 mm single-line horizontal scan protocol (length: 12 mm; depth: 2.6 mm), centred on the fovea (detected based on foveal reflex and the deepest point of internal limiting membrane from OCT) with the participant fixating on a central target. According to the manufacturer’s recommendation, images with a signal strength of 50 or higher are an acceptable signal strength limit, and any image with poor quality (unable to detect retinal, choroidal, and scleral boundary) was not considered for analysis. This ocular imaging protocol and analysis were already used in our previous publication by Dhakal et al.^21^ As the thicker choroid attenuated the OCT signals, the posterior scleral imaging was only possible in high myopes with a thinner choroid (in 25 out of 36 individuals, 70% of total high myopic eyes). Analysis was performed in 23 individuals whose retinal, choroidal and scleral boundaries were clearly visible; the rest of the participants were excluded from the study.

The spherical equivalent refraction (SER) i.e., spherical power + half of cylindrical power of all the participants, ranged from -7.81 D to -26.25 D. To compare the layer thickness between higher-degree high myopes and lower-degree high myopes, the participants were divided into <-15.00 D (N=13) and ≥-15.00 D (N=10) groups.

### Image acquisition, analysis, and measurements of scleral, choroidal and retinal thickness

Posterior retinal, choroidal and scleral thickness were determined using a custom-designed semi-automated software developed in the Centre For Technology Innovation, L V Prasad Eye Institute (for details see Dhakal et al.^21^). Right eye images were exported as current B-scan (display ratio) in a .jpg file format to obtain isotropic images that were free from stretch along vertical or horizontal directions from the instrument’s inbuilt IMAGEnet software. This format was chosen to obtain isotropic images without stretching in either the vertical or horizontal directions. The exported posterior retinal choroidal and scleral OCT images had dimensions of 1024 × 437 pixels. The experienced examiner was blinded to the refraction details of the OCT images and manually marked the retinal, choroidal, and scleral boundaries by selecting multiple sparse points that were connected to each other to form a complete boundary using cubic interpolation.^23^ The scleral boundaries were determined by taking reference from previous studies.^21, 22^ The thickness profile of all three layers was determined at the foveal and sub-foveal region, using the software (reference point chosen by the examiner). The retinal outer boundary (retino-choroidal interface) was taken as a reference boundary and corresponding normal (90 degrees from the segmenting line at retino-choroidal interface, Figure 1) to obtain retinal, choroidal and posterior scleral thickness. Retinal, choroidal and scleral thicknesses were measured as the distance between the respective inner and outer boundaries at the point of interest for each layer (Figure 1). The measurement outputs were automatically exported to a Microsoft Excel sheet (Microsoft Corporation, Redmond, WA). To assess intra-observer variability, 10 random images each of posterior retinal, choroidal and scleral thickness were analysed. The intra-observer variability was (mean ± SE) 1.23 ± 1.6 µm for retinal and choroidal thickness and 4.00 ± 6.23 µm for scleral thickness, respectively. Further detailed information on image acquisition and analysis is explained in our previous publication.^21^

**Figure. 1.**
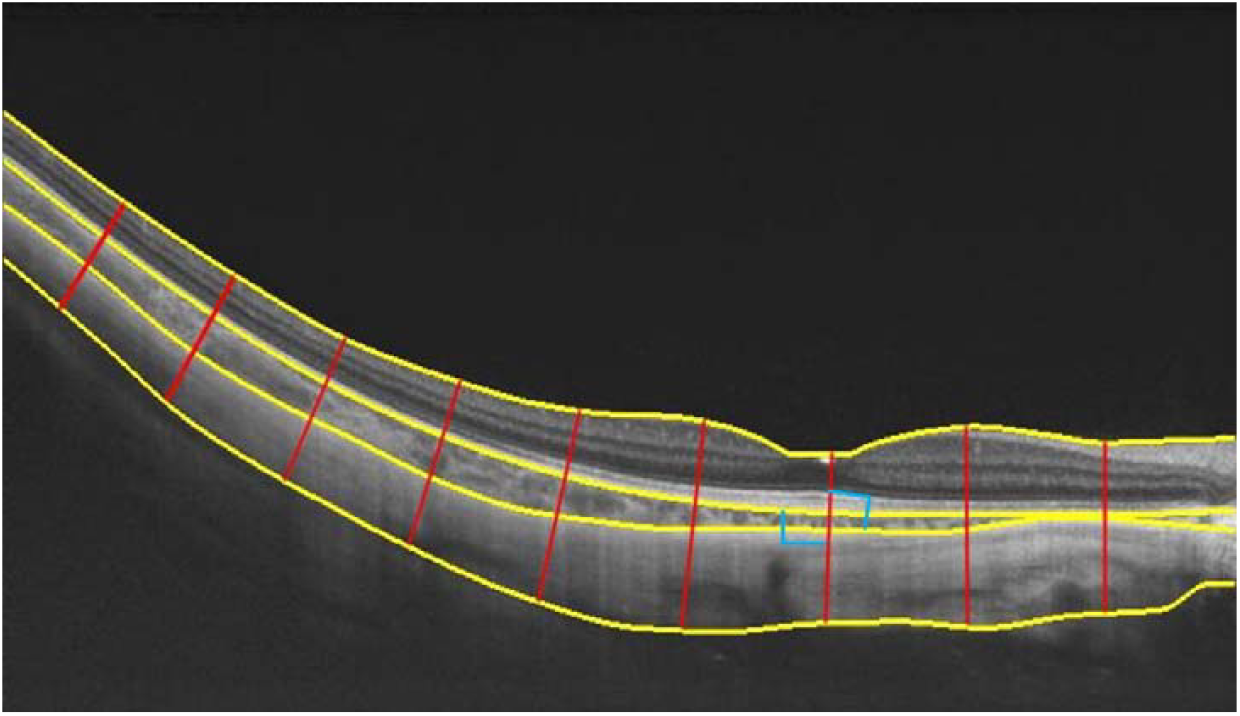
Segmented OCT image obtained from a participant with refractive error of - 23.75 D and axial length of 30.90 mm after analysis with custom-designed software, where the yellow solid lines indicate the inner and outer boundary of retina, choroid, and sclera and the red solid line is used for measuring sub-foveal retinal, choroidal, and scleral thicknesses. The angle between the segmenting line and the retino-choroidal interface was 90 degrees, as indicated with blue colour.

A sample size calculation was performed using G*Power software.^24^ Based on our hypothesis, that the inter-relationship of retinal, choroidal and scleral thickness will vary with increasing axial length, we recalculated the sample size utilizing linear multiple regression analysis, keeping 2 predictors, based on correlation coefficient values from Park et al.^16^ where, relation of axial length with choroidal and posterior scleral thickness was analysed. Based on the correlation coefficient from Park et al.^16^ we obtained sample size of 20, with an effect size f^2^ of 0.57 and a power of 80%. For the current available sample (N=23), we recalculated the study’s power using the previous effect size of 0.57, and it was 91%.

### Statistical analysis

Data were analysed using IBM SPSS Statistics 26 software (version 26, IBM Corp., Armonk, NY), and graphs were plotted using the in-built features of MS-Excel 2021 (Microsoft Corporation). Age, refractive error and ocular parameters were reported using descriptive statistics. In the Shapiro-Wilk test, the thickness of the sub-foveal retina, choroid and sclera in high myopes was normally distributed. The comparison of sub-foveal retinal, choroidal and scleral thickness was analysed with analysis of covariance (ANOVA). The relationship between axial length and the thickness of the three layers was evaluated with linear regression. The inter-relationship between retinal, choroidal and scleral thickness was evaluated with multiple regression analysis, keeping each layer and axial length as independent variables. Independent-samples t-test was used to compare between <-15.00 D (N=13) and ≥-15.00 D (N=10) groups. Statistical significance was defined as a p-value <0.05.

## Results

The mean ± SD age, spherical equivalent refractive error (SER) and axial length of all participants were 24 ± 5 years, -17.50 ± 5.32 D (Range: -7.81 to -26.25 D) and 30.17 ± 2.23 mm (Range: 27 to 36 mm), respectively. The sub-foveal retinal thickness was 245 ± 28 µm, sub-foveal choroidal thickness was 82 ± 46 µm, and sub-foveal scleral thickness was 254 ± 68 µm. The sub-foveal choroidal thickness was significantly thinner compared to the sub-foveal retina and sclera (F _(2,66)_ = 86.47, p<0.001).

With increasing axial length in high myopes, the sub-foveal choroidal thickness significantly reduced (F_1,21_ = 6.4, p=0.02); while retinal (F_1,21_ = 0.09, p=0.76), and scleral thickness variation (F_1,21_ = 0.39, p=0.54) was non-significant. With a 1 mm increase in axial length, the sub-foveal retinal thickness increased by 0.86 µm, sub-foveal choroidal thickness decreased by 10.35 µm, and sub-foveal scleral thickness decreased by 4.31 µm (Figure 2A). With decreasing spherical equivalent refraction i.e., increasing magnitude of myopia (Figure 2B), the sub-foveal choroidal thickness reduced significantly (F_1,21_ = 18.59, p<0.001). There was no significant variation in retinal (F_1,21_ = 0.36, p=0.55) and scleral thickness (F_1,21_ = 0.002, p=0.96). For a 1D myopic shift, the sub-foveal retinal thickness decreased by 0.68 µm, the sub-foveal choroidal thickness decreased by 5.88 µm, and the sub-foveal scleral thickness increased by 0.13 µm.

**Figure. 2.**
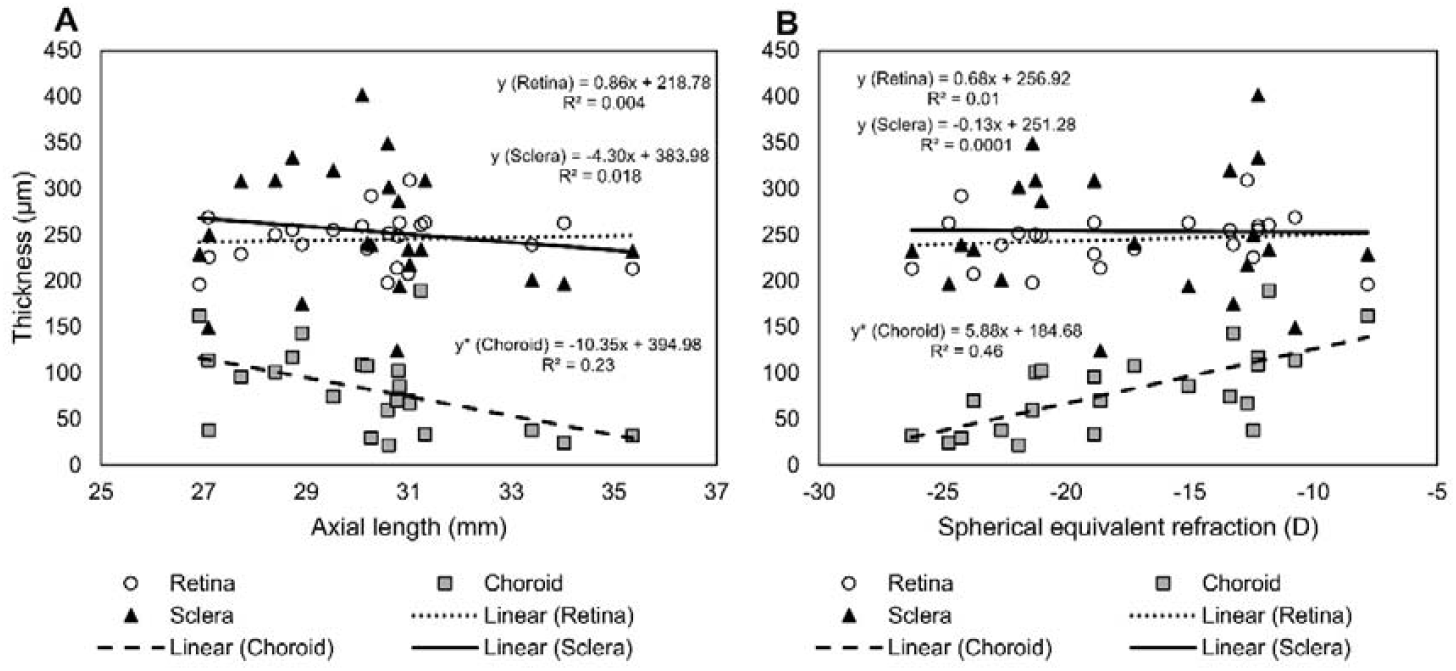
Scatter plot representing the linear regression for sub-foveal retinal, choroidal and scleral thickness with axial length for high myopes (A).Scatter plot representing the linear regression for sub-foveal retinal, choroidal and scleral thickness with spherical equivalent refraction for high myopes (B). ‘*’ Represents p<0.05.

For the inter-relationship between retinal, choroidal, and scleral thickness, the association was weak and non-significant (p≥0.10, Figure 3). With a 1 µm increase in sub-foveal choroidal thickness, the sub-foveal retinal thickness decreased by 0.05 µm, and the sub-foveal scleral thickness decreased by 0.04 µm.

**Figure. 3.**
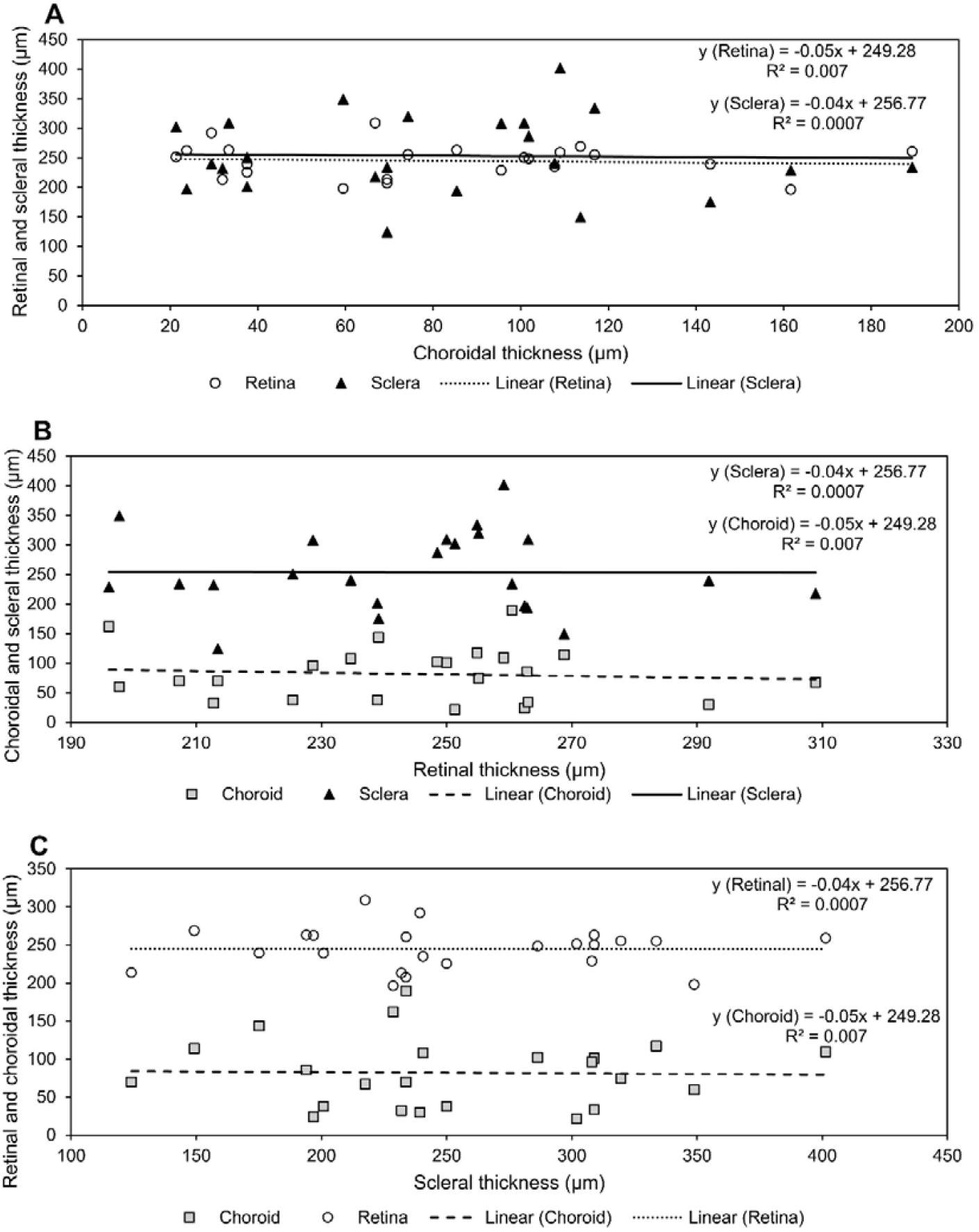
Scatter plot representing the linear regression for sub-foveal retinal and scleral thickness with sub-foveal choroidal thickness (A), sub-foveal choroidal and scleral thickness with retinal thickness (B), and sub-foveal retinal and choroidal thickness with scleral thickness (C) for high myopes. ‘*’ Represents p<0.05.

In a further sub-analysis, based on SER values, the refractive error groups were divided into <-15.00 D (N=13) and ≥-15.00 D (N=10) (Figure 4). Among the three layers, only choroidal thickness was significantly lower for the <-15.00 D group with a mean ± SD thickness of 60.16 ± 32.75 µm compared to the ≥ -15.00 D group (109.79 ± 46.10 µm, p=0.11). For retina, the thickness was 238.52 ± 26.55 µm for <-15.00 D group and 253.08 ± 29.38 µm for ≥ -15.00 D group (p=0.86). The scleral thickness was 256.20 ± 26.55 µm for <-15.00 D group and 256.20 ± 26.55 µm for ≥ -15.00 D group (p=0.52).

**Figure. 4.**
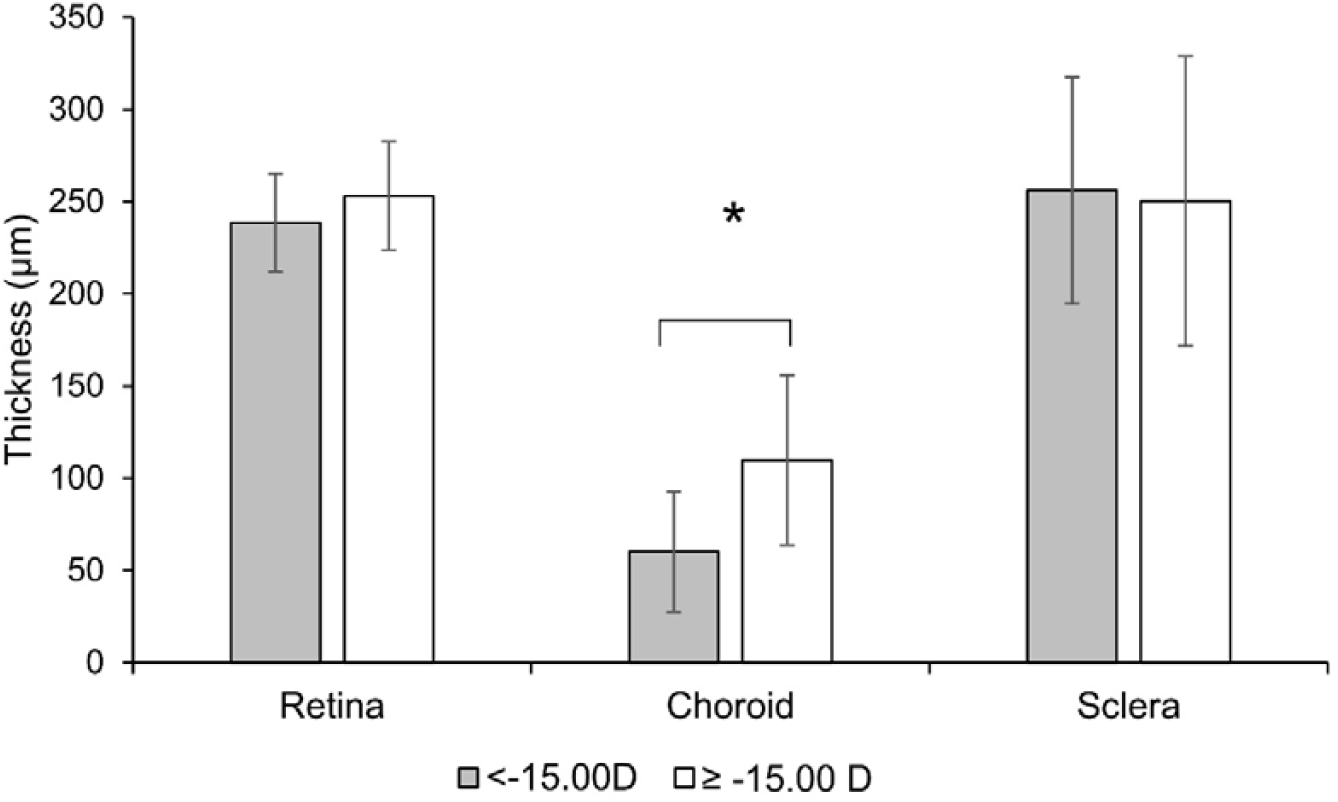
Bar plot representing the mean±SD thickness of retina, choroid and sclera for <-15.00 D and ≥ -15.00 D. ‘*’ Represents p<0.05 for comparison between two refractive error groups based on Independent-sample t-test.

## Discussion

This study investigated the sub-foveal retinal, choroidal, and scleral thickness of Indian high myopes. Three main findings from the current study are worth highlighting. First, with increasing axial length in high myopes, the sub-foveal choroidal thickness decreased significantly; however, there was no significant variation for retinal and scleral thickness. Second, with decreasing spherical equivalent refraction, only the sub-foveal choroidal thickness decreased significantly, while the sub-foveal retinal and scleral thickness variation was non-significant in high myopes. Third, the association between sub-foveal retinal, choroidal and scleral thickness was weak and non-significant.

Several studies have reported that with increasing axial length, there was no significant variation in sub-foveal retinal ^25-27^ and scleral thickness,^17-19^ while choroidal thickness decreased with increasing axial length.^28^ However, a few studies reported that the sub-foveal scleral thickness was negatively associated with axial length.^11, 16, 29^ The difference in outcomes is possibly due to the refractive error range, ethnicity, and methods used. The mean sub-foveal scleral thickness reported by Deng et al.^11^ was 491 ± 57 µm in high myopes (mean ± SD SER: −6.54 ± 1.35 D), while in our study the mean sub-foveal scleral thickness was 254 ± 68 µm (mean ± SD SER: -17.50 ± 5.32 D). The large difference in mean spherical equivalent refraction and sub-foveal scleral thickness may indicate that for larger refractive error the sub-foveal scleral thickness decreases.

For the inter-relationship between sub-foveal retinal, choroidal, and scleral thickness, the present study demonstrated findings consistent with previous reports, which have shown either no significant relation or a weak relationship between the thicknesses of these layers (Table 1).^11, 17^-^19, 29, 30^ Contrary to Park et al.,^16^ who found a significant positive relation between choroidal and scleral thickness, in the current study, there was a weak association between sub-foveal retinal, choroidal, and scleral thickness. The source of conflicting findings is unclear; differences in methodology and ethnicity could be the reason for the differences in outcomes.

**Table 1.** Compares previous studies outcomes with current study.

| Authors (year) | Population | N | SER (D) or AL(mm)<br>Mean ± SD | Age (years)<br>Mean ± SD | Instrument | AL vs RT | AL vs CT | AL vs ST | Layer vs Layer | Inference |
| --- | --- | --- | --- | --- | --- | --- | --- | --- | --- | --- |
| Maruko et al. (2012)<sup>17</sup> | Japanese | 35 | AL<br>29.0 ± 1.4 | 65.5 | SS-OCT | NA | **Slope, R<sup>2</sup>:**<br>-6.68, 0.06 | **Slope, R<sup>2</sup>:**<br>-40.10, 0.19 | **Slope, R<sup>2</sup>:**<br>CT vs ST<br>1.54, 0.19 | AL vs CT and ST: Weak association,<br>CT vs ST: Weak association. |
| Hayashi et al. (2013)<sup>19</sup> | Japanese | 54 | -12.9 ± 4.1 | 62.3 ± 11.3 | EDI-OCT | Non-significant | Non-significant | Non-significant | CT vs ST<br>Non-significant | AL vs CT and ST: No association,<br>CT vs ST: No association. |
| Park et al. (2015)<sup>18</sup> | South Korean | 24 | -7.9 ± 1.4 | 44.7 ± 10.7 | EDI-OCT | NA | NA | r: -0.10 | RNFLT vs ST<br>r: 0.46* | AL vs ST: No relation,<br>RNFLT vs ST: Significant positive relation. |
| Wong et al. (2017)<sup>29</sup> | Chinese and Malay | 41 | -12.5 ± 5.1 | 60.2 ± 8.4 | SS-OCT | NA | r: -0.44* | r: -0.42* | CT vs ST<br>r: 0.30* | AL vs CT and ST: Moderate correlation,<br>CT vs ST: Weak correlation. |
| Deng et al. (2019)<sup>11</sup> | Chinese | 810 | -2.9 ± 2.4 | 12.8 ± 3.0 | SS-OCT | **Slope, R<sup>2</sup>:**<br>4.40, 0.08* | **Slope, R<sup>2</sup>:**<br>13.0, 0.11* | **Slope, R<sup>2</sup>:**<br>-14.1, 0.09* | **Slope, R<sup>2</sup>:**<br>RT vs ST<br>0.09, 0.10*<br>CT vs ST<br>-0.18, 0.15* | AL vs RT, CT, and ST:<br>Significant association,<br>ST vs RT and CT: Significant association |
| Park et al. (2021)<sup>16</sup> | South Korean | 189 | -15.4 ± 5.4 | 63.0 ± 11.6 | WF SS-OCT | NA | ICC: -0.43* | ICC: -0.42* | CT vs ST<br>ICC: 0.28* | AL vs CT and ST: Significant negative relation,<br>CT vs ST: Significant positive relation. |
| Current study (2026) | Indian | 23 | -17.5 ± 5.3 | 24 ± 5 | EDI-OCT | **Slope, R<sup>2</sup>:**<br>0.86, 0.004 | **Slope, R<sup>2</sup>:**<br>-10.35, 0.23* | **Slope, R<sup>2</sup>:**<br>-4.30, 0.02 | **Slope, R<sup>2</sup>:**<br>RT vs CT<br>-0.05, 0.007<br>RT vs ST<br>-0.04, 0.0007<br>CT vs ST<br>-0.04, 0.0007 | AL vs CT: Significant negative association,<br>RT vs CT vs ST: Weak association. |

\*: p<0.05; SER: Spherical Equivalent Refraction; D: Diopter; N: Number of Subjects; AL: Axial length; SD: Standard Deviation; SS-OCT: Swept-Source Optical Coherence Tomography; EDI: Enhanced Depth Imaging; WF: Wide Field; R<sup>2</sup>: Coefficient of determination; ICC: Intraclass Correlation; r: Pearson correlation coefficient; RT: Retinal Thickness; CT: Choroidal Thickness; ST: Scleral Thickness; RNFLT: Retinal Nerve Fibre Layer Thickness; NA: Not Available.

Our finding of significant choroidal changes with increasing degrees of myopia, including differences between the high and very high myopic groups, but not in scleral parameters, may suggest the attainment of a saturation point for sclera, i.e. a reduced capacity of the sclera to undergo further structural alterations. However, this interpretation remains speculative at this stage, as scleral thickness measurements were not available in lower degrees of myopia for comparison in our study.

Among the three layers, only choroidal thinning was observed with increasing axial length. As reported by Wong et al.,^29^ in eyes with no or mild MMD (Myopic Macular Degeneration) the choroidal thinning was higher compared to scleral thinning with increasing axial length, which could be a reason for only choroidal thinning with axial length in the current study.

Various theories were proposed to indicate the choroid as a marker for ocular elongation.^2, 4, 31, 32^ These theories have emphasized the influence of choroidal thickness and vasculature on scleral remodeling. Based on the week relation between retinal, choroidal and scleral thickness, we speculate that in high myopes during axial elongation, the choroidal structural changes may not directly influence the thickness of the posterior sclera, and other factors, such as alteration in biochemical signaling and scleral molecular structure, play a contributing role whose understanding is beyond the scope of this manuscript.

This is the first study to report the relationship among retinal, choroidal, and scleral thickness at sub-foveal regions in Indian high myopes. One of the major limitations of our study is that we could not acquire the scleral thickness of emmetropes and low myopes due to instrumental limitations. Further studies with the advancement of imaging technology can be conducted involving emmetropes and low myopes to understand the possible relationship between retinal, choroidal and scleral thickness in the non-foveal region. As it is difficult to get non-pathologic high myopes with axial length >33 mm, we had only 3 participants; more myopes with higher axial length could have strengthened the results in the current study. Given the cross-sectional nature of the current study, further longitudinal studies are warranted to understand the interaction among the retinal, choroidal and scleral thickness with myopia progression in farther periphery.

In conclusion, with increasing axial length and severity of myopia in high myopes, compared to scleral and retinal thickness, the sub-foveal choroidal thickness decreased significantly. The association among sub-foveal retinal, choroidal, and scleral thickness was weak and non-significant, which indicates that all the outer coats of the eye ball do not necessarily stretch in equal magnitude. Changes in choroidal thickness do not necessarily reflect the changes in retinal and scleral thickness.

The findings from our study involving high myopes re-emphasize the potential role of choroid as a marker for ocular elongation and add speculation that the sclera may have reached a limit with ocular elongation, beyond which further thinning is unlikely with increasing axial length, which needs to be investigated further in a large sample size.

## Data Availability

All data produced in the present study are available upon reasonable request to the authors

## Acknowledgement

The authors thank Hyderabad Eye Research Foundation, L V Prasad Eye Institute, India for the support in conducting the study.

## Conflict of interest

None for SP, RD and KKV. PKV has received funding from SEED, Nthalmic, EssilorLuxottica and filed patents for Peripheral-Ancillary Refractive Component (PARC) and Myopia predictive technology. None of these fundings were contributed to the current study.

**All co-authors vouch for the accuracy of the data and agree to have read and approved the final draft of the manuscript**

## Data availability statement

The data that support the findings of this study are available on request from the corresponding author, [PKV]. The data are not publicly available due to their containing information that could compromise the privacy of research participants.

